# LM-QASAS: Reference-free identification of antigen-specific sequences from the BCR repertoire using antibody language models

**DOI:** 10.64898/2026.03.31.26349834

**Authors:** Genki Masuda, Yohei Funakoshi, Shunsuke Iizumi, Kimikazu Yakushijin, Goh Ohji, Hironobu Minami, Masahito Ohue

## Abstract

The B-cell receptor (BCR) repertoire serves as a historical record of immunological events. However, deciphering antigen-specific sequences from this vast dataset remains a challenge, particularly for novel pathogens where prior knowledge is absent. While time-course analysis methods such as QASAS have proven effective for tracking immune responses, they rely on existing antibody databases, limiting their applicability to emerging diseases. To overcome this limitation, we introduce LM-QASAS, a reference-free computational framework that integrates antibody language models with repertoire dynamics. By mapping sequences into a high-dimensional semantic embedding space, LM-QASAS identifies functionally convergent clusters of sequences that are semantically similar and exhibit transient expansion upon immune stimulation. In healthy individuals vaccinated with SARS-CoV-2 mRNA vaccines, our method identified spike-specific sequences with over 90% purity, significantly outperforming methods based on simple sequence identity or abundance. Leave-one-out cross-validation demonstrated that LM-QASAS could accurately reconstruct immune dynamics in unseen individuals without external references. Conversely, the method showed limited sensitivity in an influenza vaccine cohort, revealing that the approach is most effective under conditions of robust clonal expansion (high signal-to-noise ratio), such as those induced by mRNA vaccines. LM-QASAS provides a rapid, high-precision platform for monitoring humoral immunity against emerging threats.

**Significance statement:** Antibodies that recognize a pathogen leave traces in a person’s B-cell receptor (BCR) repertoire, but identifying those antigen-specific sequences is difficult, especially for new diseases lacking reference databases. We present LM-QASAS, a reference-free method that combines antibody language models with time-course repertoire data to detect clusters of sequences that expand transiently after immune stimulation. In SARS-CoV-2 mRNA vaccinees, LM-QASAS enriched spike-related sequences with very high precision and enabled reconstruction of immune-response dynamics in unseen individuals using pseudo-reference databases built from other subjects. Analyses of convalescent, post-transplant, and influenza cohorts clarify when the approach works best: strong clonal expansion and a high signal-to-noise ratio. This framework supports rapid monitoring of humoral immunity against emerging threats.

## Introduction

The adaptive immune system generates a theoretical diversity of up to 10^14^ variants via B-cell receptor (BCR) gene rearrangement, thereby addressing a vast range of pathogens^1, 2^. The BCR repertoire accumulated within an individual reflects their immunological history, including infections, vaccinations, and autoimmune responses, and serves as a rich source of information for comprehensively assessing the state of the adaptive immune system^3, 4^. Advances in next-generation sequencing (NGS) technologies have enabled the exhaustive analysis of this massive information, which became a key to elucidating immune dynamics during the COVID-19 pandemic^5–9^. However, deciphering definitive antibody sequences related to specific diseases from these vast datasets without prior knowledge remains a significant challenge.

Significant progress has been made in tracking antigen-specific responses from longitudinal BCR repertoire data in recent years. For instance, the Quantification of Antigen-Specific Antibody Sequence (QASAS) method proposed by Funakoshi et al. is a powerful framework for quantitatively visualizing responses to specific antigens by referencing known antibody sequence databases^10, 11^. However, this approach relies on the existence of referenceable antibody databases. With exceptions like COVID-19, such databases are largely undeveloped for emerging infectious diseases and many intractable conditions. Thus, to realize truly versatile repertoire analysis, a technology that autonomously discovers antigen-specific sequences from the internal structure and dynamics of the repertoire data itself, without relying on external ground truth data, is indispensable. To date, experimental approaches such as LIBRA-seq and computational methods based on sequence similarity or convergence have been proposed to identify antigen-specific BCRs^12–15^. In particular, approaches focusing on identical amino acid sequences shared among individuals (public clones) are effective as a baseline for tracking strong immune responses. However, experimental methods face challenges regarding cost and throughput, while sequence identity-based methods have limitations: they risk missing clones that are diverse at the sequence level but functionally equivalent, and they cannot fully exclude non-specific sequence contamination. Furthermore, recently reported specificity prediction using large language models often relies on supervised learning^16^, leaving challenges for immediate application to unknown antigens.

How can functional immune signals be extracted with high purity in the absence of known ground truth? To address this challenge, we introduce a semantic approach using protein language models^17^. An antibody language model (AbLM), pre-trained on massive antibody sequences, embeds amino acid sequences as high-dimensional vectors similar to natural language processing, capturing biological context and structural features beyond simple sequence identity^18, 19^. In this semantic space, functionally convergent clones are expected to be placed in proximity even if they are distant at the sequence level, enabling the detection of antigen-specific populations that could not be captured by conventional sequence analysis.

In this study, we propose LM-QASAS (Language Model-guided QASAS), a novel reference-free computational framework that integrates AbLM embeddings with BCR repertoire temporal dynamics. The core of LM-QASAS lies in mapping antibody sequences to semantic space via AbLM and detecting temporal fluctuations in local density within that space. Specifically, we employ two complementary approaches, discrete clustering (*K*-means)^20^ and continuous kernel density estimation (KDE)^21–23^, to identify semantic clonal populations that selectively expand following interventions such as vaccination. This enables the high-precision extraction of antigen-specific antibody candidates without any reliance on external databases.

We validated LM-QASAS using longitudinal BCR repertoire data from COVID-19 vaccinees, demonstrating that the method can identify SARS-CoV-2 spike-specific antibody sequence candidates without prior knowledge. In particular, we reveal that LM-QASAS can enrich antigen-specific sequences with extremely high purity under strong immune stimulation compared to methods based on simple sequence convergence. Furthermore, we demonstrate that QASAS analysis using the extracted sequences allows for the precise quantification of the dynamics of the specific response’s onset and convergence. LM-QASAS serves as a powerful computational framework for monitoring humoral immune responses in scenarios where reference data are absent and robust immune responses are expected, such as rapid vaccine evaluation for emerging infectious diseases.

## Results

### Overview of LM-QASAS and semantic-space immune dynamics

Figure 1 illustrates the analytical workflows of the existing QASAS method, which quantifies antigen-specific responses from longitudinal BCR repertoires, and our proposed LM-QASAS, a reference-database-independent framework. QASAS visualizes immune responses by cross-referencing BCR sequences obtained pre- and post-vaccination against public neutralizing antibody databases and tracking the temporal changes in the hit rate of sequences similar to database entries (Fig. 1a). In contrast, LM-QASAS extracts sequence populations that exhibit transient expansion at the peak time point within the embedding space of an antibody language model, based solely on the internal dynamics of the repertoire, thereby constructing a pseudo-database for unknown antigens (Fig. 1b). While Fig. 1b shows an example that uses kernel density estimation on UMAP projections for visual clarity, our actual analysis evaluated both a KDE-based approach and a *K*-means-based approach. The latter partitions the original high-dimensional embedding space to identify candidates based on temporal fluctuations of clusters, allowing a comparative assessment of the two variants.

**Figure 1.**
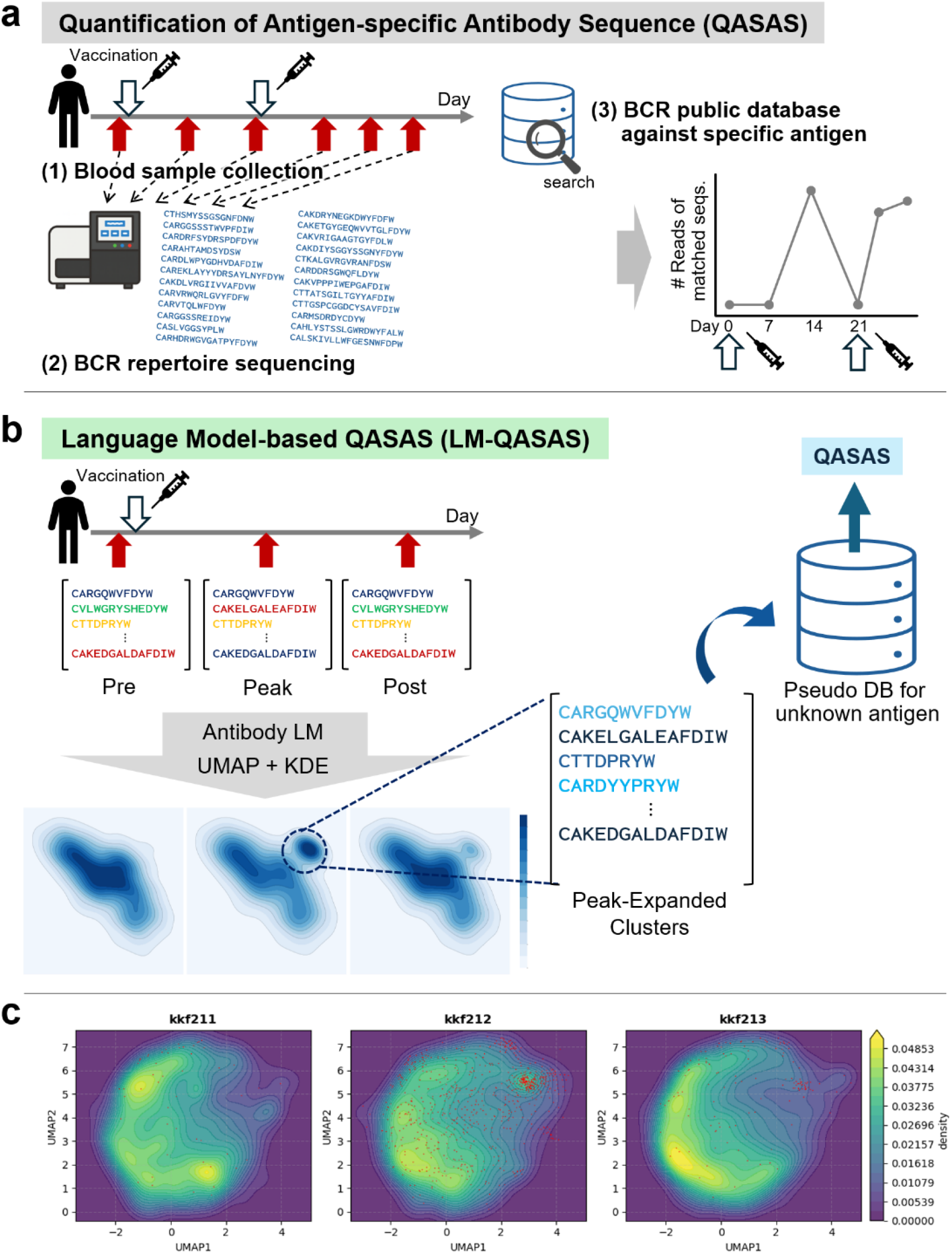
Overview of QASAS and the proposed reference-free LM-QASAS, and an example of semantic-space repertoire dynamics. (a) QASAS workflow. Blood samples are collected before and after SARS-CoV-2 mRNA vaccination to obtain BCR repertoire sequences. These are cross-referenced with a public neutralizing antibody database to quantify immune response dynamics based on the rise in the hit rate of sequences similar to database entries at the peak time point. (b) LM-QASAS workflow. Longitudinal repertoire sequences are converted into embeddings using an antibody language model. Densities at each time point are estimated via KDE in a UMAP-projected space. Expanded sequence clusters are extracted from regions exhibiting increased local density at the peak followed by a decline. These clusters form a pseudo-database for unknown antigens, enabling time-series analysis equivalent to QASAS without reliance on external references. (c) Representative semantic-space visualization. The BCR repertoire of a representative healthy mRNA vaccinee is projected into 2D space using UMAP based on AbLM embeddings, with distribution densities visualized via KDE for three time points. Red points indicate sequences similar to those in a known SARS-CoV-2 antibody database.

### Characterization of semantic space and cluster dynamics

The LM-QASAS framework proposed in this study is grounded in the hypothesis that antigen-specific immune responses manifest as increases in local sequence density within the semantic space constructed by an AbLM. To validate the biological validity of this hypothesis, we analyzed longitudinal BCR repertoire data from three cohorts with distinct immune backgrounds: healthy SARS-CoV-2 mRNA vaccinees, COVID-19 convalescent patients, and post-hematopoietic stem cell transplantation mRNA vaccinees.

First, to visually demonstrate the fundamental principle of our method, we selected a representative subject from the healthy vaccinee cohort and analyzed the spatiotemporal dynamics of their repertoire in detail. Specifically, we vectorized all BCR sequences at three time points, pre-vaccination, the peak of antibody titer rise, and the subsequent decline phase, using AbLM and projected them into a two-dimensional space using UMAP. Furthermore, we visualized the density of the sequence distribution at each time point using kernel density estimation (Fig. 1c). To interpret the biological meaning of this spatial arrangement, we defined sequences within the subject’s repertoire that exhibited high similarity (Levenshtein distance ≤ 2) to those registered in the known SARS-CoV-2 neutralizing antibody database, CoV-AbDab^11^, as known antigen-specific sequences and plotted them onto the same space (Fig. 1c, red points).

The analysis revealed that the BCR repertoire was widely dispersed throughout the semantic space at all three time points, confirming the maintenance of a diverse antibody population. However, focusing on the local density distribution revealed dramatic changes. At the pre-vaccination stage, CoV-AbDab-related sequences were barely detectable, and no significant concentration in specific regions was observed. In contrast, at the peak of the immune response, a clear increase in density was confirmed in a specific local region that had been low-density prior to vaccination. Crucially, the CoV-AbDab-related sequences that appeared in large numbers at this timing were distributed extremely intensively within this newly formed high-density region. Subsequently, over time, the density of the region decreased and the number of red points declined, but we observed that a subset of sequences was retained in the same region as the peak.

This visual evidence demonstrates that within the AbLM-based semantic space, antigen-specific clonal expansion is identifiable as a local elevation in specific semantic regions distinct from the diverse background repertoire. It strongly suggests that, by tracking changes in high-density regions, it is possible to trace the dynamics of antigen-specific antibody populations from emergence to convergence without prior reference to external databases.

### Validation with known neutralizing antibody databases

The LM-QASAS framework proposed in this study is designed to extract high-density clusters that form locally within semantic space, as observed in the previous section. In this section, we evaluated two variants of the cluster extraction approach: LM-QASAS (*K*-means), based on discrete clustering, and LM-QASAS (KDE), based on continuous density estimation.

To validate the antigen specificity of the extracted sequences, we analyzed the top 300 candidate sequences extracted from each subject and calculated the proportion of sequences containing high similarity (Levenshtein distance ≤2) to entries registered in CoV-AbDab. We established the following three comparative methods.

- Reads: top sequences based on read counts at the peak of the immune response.
- Random: sequences randomly sampled from the repertoire at the peak time point.
- Dups: sequences with different V/J gene combinations but identical CDRH3 amino acid sequences, extracted based on their diversity count. This serves as a baseline for evaluating public clones that converge among individuals.

The analysis included a total of 10 subjects across three cohorts with distinct immune backgrounds: healthy mRNA vaccinees, COVID-19 convalescent patients, and post-HSCT mRNA vaccinees. The evaluation results for each subject are presented in Fig. 2.

**Figure 2.**
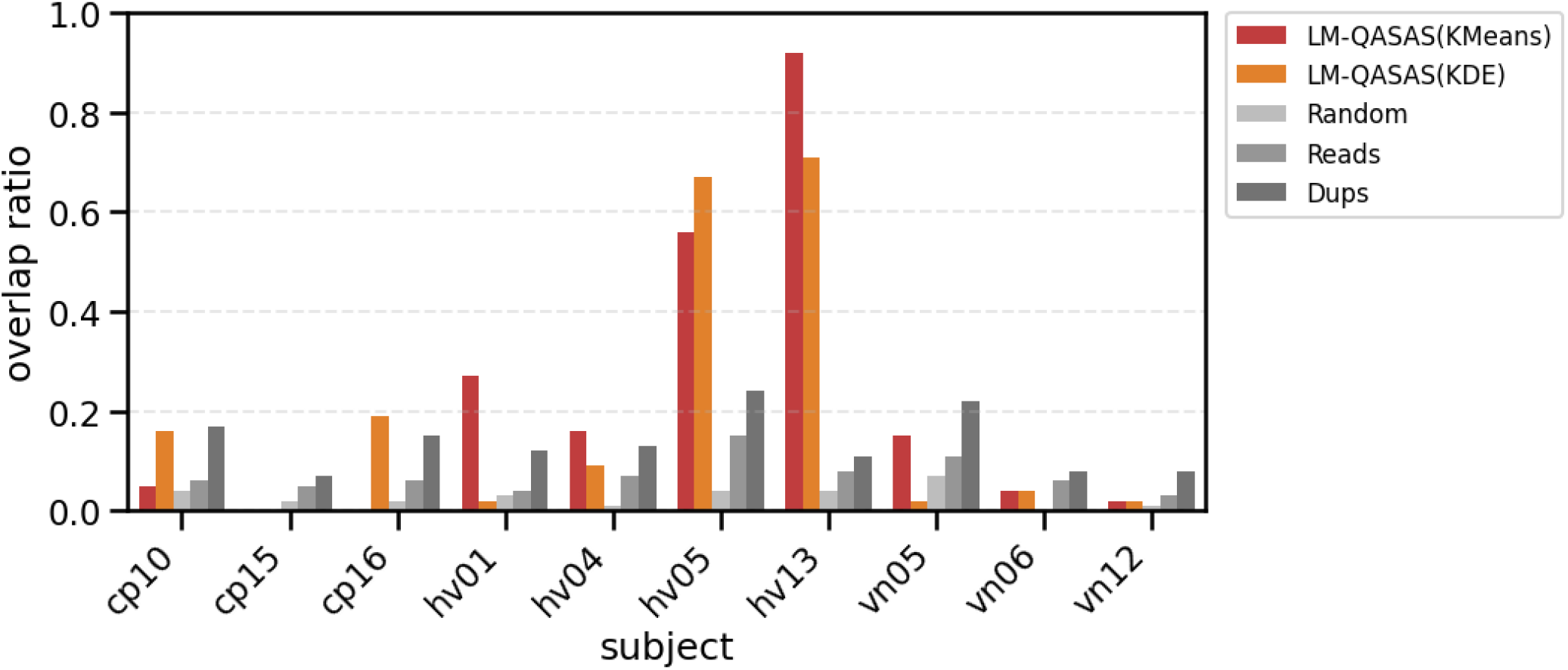
Comparison of the proportion of CoV-AbDab-related sequences across subjects and cohorts. Proportion (%) of CoV-AbDab-related sequences within the top 300 sequences extracted from 12 subjects. The prefixes in subject IDs denote healthy vaccinees, convalescent patients, and post-HSCT vaccinees. In the healthy vaccinee group, both LM-QASAS variants exhibited high proportions, outperforming Dups. In the convalescent and post-HSCT groups, the performance of the *K*-means variant declined; however, the KDE variant achieved proportions comparable to Dups in a subset of the convalescent group.

In the healthy vaccinee group, both LM-QASAS variants and Dups exhibited a higher proportion of CoV-AbDab-related sequences compared to Reads and Random. Notably, the performance of LM-QASAS was superior, with both the *K*-means and KDE variants achieving higher proportions than Dups. In certain subjects, such as HV13, over 90% of the sequences extracted by LM-QASAS were CoV-AbDab-related. This confirms that under conditions of strong antigen stimulation, our method can enrich antigen-specific sequences with extremely high purity.

Conversely, in the convalescent and post-transplant groups, distinct behaviors were observed among the methods. The proportion of validated sequences for LM-QASAS (*K*-means) dropped significantly in these groups, falling to levels comparable to Random in many subjects. In contrast, Dups consistently maintained higher proportions than Random or Reads even in the convalescent and post-transplant groups, demonstrating robustness in detecting public clones.

Interestingly, LM-QASAS (KDE) exhibited a different behavioral pattern compared to *K*-means. While performance remained low in the post-transplant group, similar to *K*-means, the KDE variant achieved a proportion of approximately 20% in two out of three subjects in the convalescent group, demonstrating performance comparable to Dups. Immune responses induced by natural infection target a more diverse array of antigens compared to vaccination, and the resulting cluster shapes in the semantic space are expected to be heterogeneous. This result suggests that the KDE method, which captures continuous gradients, may be more sensitive to the complex response patterns derived from natural infection than the *K*-means method, which relies on discrete centroids.

These results indicate that while LM-QASAS, particularly the *K*-means variant, demonstrates overwhelming specificity in healthy vaccine responses, for more complex infection-induced responses, the KDE approach or convergence analysis methods like Dups may function in a complementary manner.

### Tracking immune dynamics in unseen individuals

Finally, to demonstrate the generalizability of our method to unseen individuals, we evaluated the time-series tracking performance using leave-one-out cross-validation. In this experiment, we used LM-QASAS (*K*-means) to extract the top 300 antigen-specific candidate sequences from 9 out of the 10 subjects. By integrating these, we constructed a pseudo-reference database consisting of a total of 2,700 sequences. We then performed QASAS analysis on the longitudinal repertoire data of the remaining one subject using this pseudo-database to quantify the dynamics of the immune response. This process was repeated for all subjects to verify whether immune dynamics could be accurately tracked without relying on external databases.

As comparators, we used CoV-AbDab as the ideal ground truth and a randomly sampled sequence group as a baseline. For the Random group, we similarly extracted 300 sequences randomly from the peak-time repertoire of each subject to create a database of 2,700 sequences. It should be noted that the number of unique CDRH3 amino acid sequences in CoV-AbDab is about 4.5 times larger than the database size of our method and the Random group; therefore, the scale of the absolute values of the calculated overlapping rates differs.

The visualization results of the time-series trends for all subjects are shown in Fig. 3. The time-series profiles using sequences extracted by LM-QASAS exhibited immune response dynamics extremely similar to the profiles using CoV-AbDab. Generally, it is known that the peak of the humoral immune response appears around 14 days after the event for infected individuals and primary vaccinations, and around 7 days after vaccination for second or subsequent doses involving memory B cells^24^. Consistent with this biological knowledge, the LM-QASAS profiles successfully and accurately captured the rapid score increase, peak timing, and subsequent convergence in each cohort. Particularly in the healthy vaccinee group, compared to the Random group using a database of the same size, it was confirmed that peak detection was achieved with greater sensitivity and higher signal intensity.

**Figure 3.**
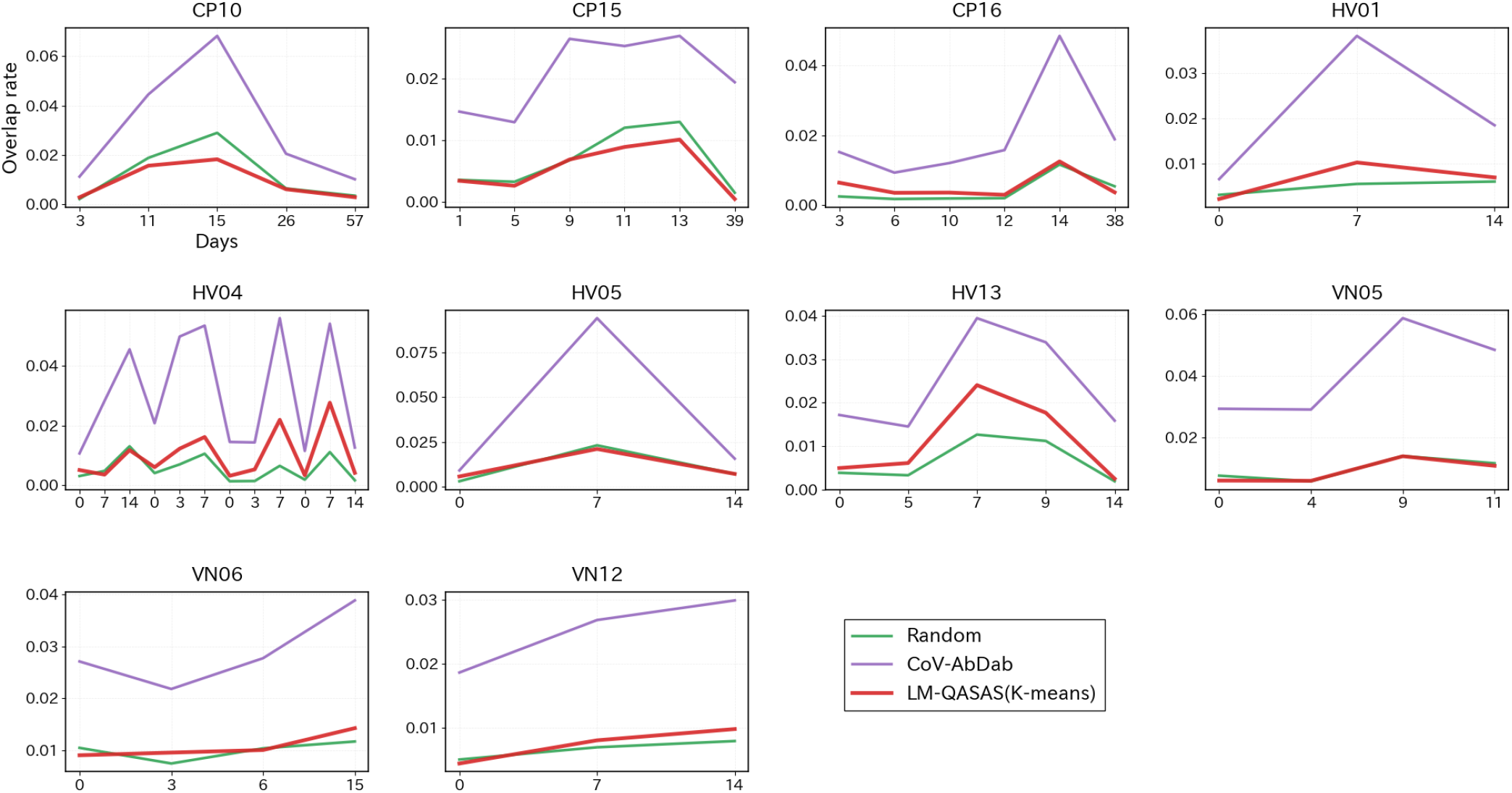
Visualization of immune response dynamics using leave-one-out cross-validation. Results of QASAS analysis performed using a database of sequences extracted by LM-QASAS from other subjects’ data in a leave-one-out cross-validation setting. The trajectory of LM-QASAS shows an immune response pattern similar to that of the ground truth CoV-AbDab, accurately capturing cohort-dependent peak timing. Particularly in the healthy vaccinee group, LM-QASAS forms distinct peaks compared to Random using a database of the same scale.

### Evaluation on influenza vaccine cohort

To assess the applicability and limitations of LM-QASAS, we performed a similar leave-one-out cross-validation on an influenza vaccine cohort (*n* = 17), which presents a different antigen and immune background from SARS-CoV-2. In this experiment, we utilized repertoire data from three time points: pre-vaccination (Day 0), post-vaccination (Day 7), and two weeks post-vaccination (Day 14).

The analysis results for four representative subjects are shown in Fig. 4. In contrast to the clear tracking accuracy observed in the SARS-CoV-2 cohort, in the influenza cohort, immune response dynamics could not be clearly visualized in 11 out of 17 subjects, even when using LM-QASAS. While an increase in the overlapping rate at the peak was confirmed in some subjects, distinguishing the signal from the baseline proved difficult in the majority of cases.

**Figure 4.**
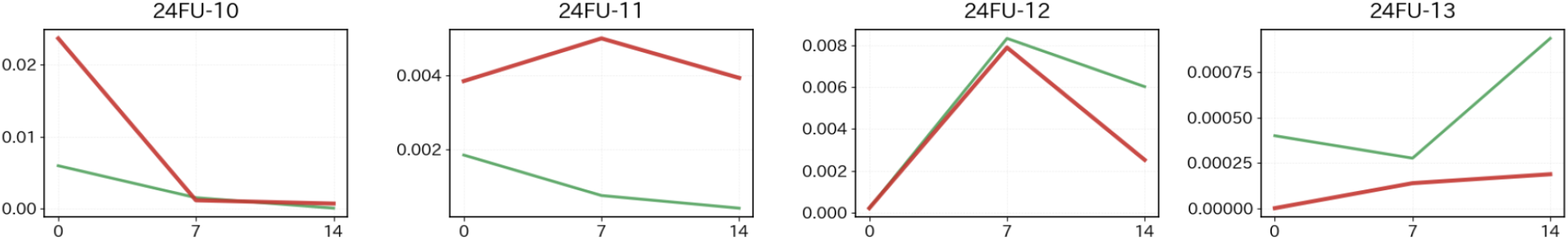
Analysis of immune response dynamics in influenza vaccinees. Profiles of four representative subjects from the leave-one-out cross-validation performed on the influenza vaccine cohort. Unlike the SARS-CoV-2 cohort, peak detection by LM-QASAS was unclear in many subjects. The nearly flat trajectory of Random suggests that the compositional change in the entire repertoire due to influenza vaccination is minimal.

Furthermore, we compared the trends of the Random extraction group to assess global repertoire fluctuations. While the SARS-CoV-2 cohort showed a clear rise at the peak, the influenza cohort exhibited an almost flat trend across all time points. We consider the behavior of the Random group to reflect the proportion of antigen-specific clones within the entire repertoire, that is, the signal-to-noise ratio. Therefore, these results indicate that repertoire fluctuations in peripheral blood induced by influenza vaccination are extremely limited compared to those induced by mRNA vaccination. Since LM-QASAS relies on detecting density changes, it was revealed that the detection sensitivity for antigen-specific signals decreases under conditions where global repertoire fluctuation is scarce, namely when the signal-to-noise ratio is low.

## Discussion

In this study, we proposed LM-QASAS, a method for identifying antigen-specific B-cell receptors without reliance on known databases, by analyzing the density dynamics of antibody sequences within a semantic space constructed by an antibody language model. Comprehensive validation using COVID-19 and influenza vaccine cohorts revealed that while this method can enrich antigen-specific sequences with extremely high specificity under conditions of robust immune stimulation, it exhibits detection limits in recall responses where global repertoire fluctuations are minimal.

A key finding of this study is that the detection performance of LM-QASAS varied markedly depending on subjects’ immune backgrounds. These variations reflect the qualitative and quantitative differences in immune responses across the cohorts.

First, the high accuracy achieved by LM-QASAS in healthy vaccinees can be attributed to the specific characteristics of mRNA vaccines. Previous studies have shown that, compared to conventional protein vaccines, mRNA vaccines induce more robust and persistent germinal center reactions against a single antigen and cause massive mobilization of plasmablasts^25, 26^. This synchronous and explosive clonal expansion likely results in the formation of high-density clusters in specific regions of the semantic space, allowing the density-based extraction algorithm to function ideally.

In contrast, the reduced scores observed in convalescent patients are likely related to differences in target antigens. While mRNA vaccines target only the spike protein, natural infection exposes the immune system to the whole virus particle, inducing a broad polyclonal response not only against the spike protein but also against internal viral components such as the nucleocapsid and membrane proteins^27, 28^. However, the CoV-AbDab database referenced in this study is biased toward spike antibodies. Consequently, even if LM-QASAS successfully extracted genuine antibody clusters specific to infection, such as those targeting non-spike antigens, they were likely classified as mismatches in this validation.

Furthermore, the lower performance in post-hematopoietic stem cell transplantation patients may reflect the structural instability of the repertoire during immune reconstitution. Post-transplant B-cell repertoires are known to exhibit lower diversity compared to healthy individuals due to insufficient supply of naive B cells and homeostatic proliferation^29, 30^. Under such conditions, vaccine-specific clonal expansion is easily buried in background noise, which likely made signal extraction via density estimation difficult. However, given that the Dups method achieved a certain level of success, it is evident that the mobilization of public clones is occurring, suggesting a need for future algorithmic improvements to amplify weak signals in such complex backgrounds.

The discrepancy in results between the SARS-CoV-2 and influenza cohorts highlights the immunological conditions required for this method to function effectively, particularly the importance of the magnitude of induced clonal expansion. The SARS-CoV-2 cohort comprised many multi-dose vaccinees, and recall responses mobilizing existing memory B cells are also expected. However, a decisive difference was observed in repertoire fluctuations between the two groups. It is known that mRNA vaccines, due to sustained intracellular antigen production and high immunogenicity, induce extremely large-scale plasmablast bursts detectable in peripheral blood and persistent germinal center reactions, even during recall responses^25, 26^. This potent induction resulted in a drastic increase in the proportion of antigen-specific clones within the total repertoire, likely allowing the density detection mechanism of LM-QASAS to function effectively.

In contrast, seasonal influenza vaccination in adults often elicits a more modest magnitude of *de novo* clonal expansion in peripheral blood compared to mRNA vaccines^31, 32^. Although the recall response induced by influenza vaccines is rapid and specific, it often does not involve a substantial physical increase in sequence numbers sufficient to alter the global structure of the repertoire. Consequently, the signal-to-noise ratio in the influenza cohort remained low, and the density detection sensitivity of LM-QASAS was insufficient to capture the signal.

These comparisons suggest that, regardless of immune history, this method is best suited for analyzing events where robust clonal expansion is induced by strong immune stimulation, such as that provided by mRNA vaccines.

## Conclusion

This study demonstrates that LM-QASAS is a powerful computational framework capable of identifying antigen-specific antibodies with high purity and precision, independent of external reference databases.

Our comparative analysis revealed a critical hierarchy for extracting genuine immune signals from repertoire data. First, approaches focusing on inter-individual convergence, such as Dups and LM-QASAS, captured antigen-specific sequences more reliably than analyses based on simple abundance. This suggests that genuine immune responses are more robustly represented as shared population-level patterns than as highly expanded private clones derived from individual immune history.

Second, by incorporating antibody language models, LM-QASAS extends the detection of convergence from exact sequence identity to functional similarity in semantic space. By capturing functional convergence beyond sequence-level differences, LM-QASAS reduces noise that simple sequence identity-based methods cannot exclude, and achieves very high sequence enrichment under conditions of strong immune stimulation.

Although the current framework has limitations in sensitivity under low signal-to-noise conditions, it provides a versatile and responsive strategy for repertoire analysis in settings where external reference data are unavailable. In particular, LM-QASAS may support rapid vaccine evaluation and immune-response monitoring during the early phase of emerging infectious diseases.

## Methods

### Study datasets and ethics

This study was approved by the Medical Research Ethics Committee of the Kobe University Hospital Ethics Committee (Protocol no. B240101) and was conducted in accordance with the Declaration of Helsinki and relevant ethical guidelines. All samples were collected at Kobe University Hospital between July 2020 and January 2025. Written informed consent was obtained from all participants prior to participation.

For the analysis, we used longitudinal BCR repertoire data from SARS-CoV-2-related cohorts obtained in a previous study by Funakoshi et al.^10^, as well as separately obtained data from an influenza vaccine cohort. The breakdown of the SARS-CoV-2-related cohorts, as shown in Table 1, consists of COVID-19 convalescent patients (*n* = 3), healthy volunteers vaccinated with mRNA vaccine (Pfizer BNT162b2) (*n* = 4), and patients vaccinated with the same vaccine after hematopoietic stem cell transplantation (*n* = 3). Peripheral blood samples were collected longitudinally from each subject for up to two months, centering around two weeks after infection or vaccination. For the influenza vaccine cohort, peripheral blood samples were obtained from 17 healthy individuals vaccinated with seasonal influenza vaccine at pre-vaccination (Day 0), 7 days post-vaccination (Day 7), and 14 days post-vaccination (Day 14).

**Table 1.** Details of the SARS-CoV-2 cohorts used in this study.

| Group ID | Description | *n* | Sampling Period |
| --- | --- | --- | --- |
| **CP** | COVID-19 Convalescent Patients Infected with SARS-CoV-2 | 3 | Aug. – Dec. 2020 |
| **HV** | Healthy Volunteers Vaccinated with mRNA BNT162b2, Naive to SARS-CoV-2 | 4 | May – Dec. 2021 |
| **VN** | Vaccinated Patients post-Hematopoietic Stem Cell Transplantation | 3 | Dec. 2022 – Jan. 2024 |

### BCR library preparation and sequencing

BCR repertoire analysis was performed using unbiased next-generation sequencing technology developed by Repertoire Genesis Inc.^33^. Analysis of the SARS-CoV-2-related cohorts was performed by Repertoire Genesis Inc., while analysis of the influenza vaccine cohort was performed by Takara Bio Inc.; however, the basic experimental protocols and analysis pipelines are common.

Specifically, cDNA was synthesized from total RNA using SuperScript III Reverse Transcriptase (Invitrogen) and polyT18 primers. After synthesizing double-stranded cDNA, a dsDNA adaptor was ligated and digested with NotI restriction enzyme. Subsequently, nested PCR was performed using KAPA HiFi DNA Polymerase with IgG constant region-specific primers and an adaptor primer. Amplicon libraries were prepared by a second PCR amplification, and index sequences were added using the Nextera XT Index Kit v2 Set A. Sequencing was performed using the Illumina MiSeq paired-end platform (2 × 300 bp).

### Sequence data processing

Repertoire analysis software proprietary developed by Repertoire Genesis Inc. was used for processing the obtained sequence data. Each sequence was assigned V, D, and J genes and CDRH3 regions were identified based on identity with reference sequences from the international ImMunoGeneTics information system database^34^. Strict filtering criteria were applied to ensure the reliability of the analysis. Specifically, only sequences containing no stop codons and that were in-frame were retained. In addition, only sequences with functional V, D, and J genes were included in the analysis; sequences determined to be ORFs or pseudogenes were excluded. Furthermore, sequences with a CDRH3 amino acid length of less than 5 were excluded from the analysis to eliminate alignment errors and non-specific fragments.

### Definition of unique clones and input features

In this study, sequence groups in which all four elements, V gene, J gene, CDRH3 amino acid sequence, and isotype, matched were defined as the same BCR clone. Therefore, even if the CDRH3 amino acid sequences are identical, they are treated as separate unique clones if other elements such as V gene or isotype differ. In the LM-QASAS analysis, each of these unique clones is treated as a single input data point. For the calculation of embedding representations by AbLM, only the CDRH3 amino acid sequence of each clone is used; V/J gene and isotype information are not used for vector generation. Furthermore, since this method focuses on the density distribution of sequences in the semantic space, read count information for each unique clone was intentionally excluded from the analysis. By treating all unique clones as equivalent points, this approach eliminates bias caused by high-frequency clones and aims to detect variants that are functionally important despite their low frequency.

### LM-QASAS algorithm

LM-QASAS is a method that identifies antigen-specific clusters based on density fluctuations in semantic space. In this study, we extracted sequence groups showing transient expansion and contraction associated with vaccination or infection using the following scoring functions.

### LM-QASAS (K-means)

For each subject, unique sequences from all time points were integrated, and the embedding vectors were divided into *K* clusters (here *K* = 100) using the *K*-means method. For each cluster *C*_*i*_, the following score integrating the rate of increase from pre-immune response to peak and the rate of decrease from peak to post-immune response was calculated:

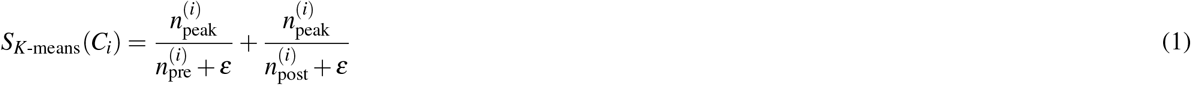

where 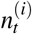 is the number of sequences in cluster *C*_*i*_ at time point *t*, and *ε* is a pseudo-count to prevent division by zero. Finally, among the sequences contained in the repertoire at the peak time point, sequences belonging to clusters with high scores were extracted as antigen-specific candidates.

### LM-QASAS (KDE)

To evaluate the local density of sequences in more detail, first, all sequences for each subject were projected into a two-dimensional space using UMAP. In this two-dimensional space, kernel density estimation was performed for samples at each time point *t* to estimate the probability density function *d*_*t*_ (*x*) at an arbitrary coordinate *x* within the space. The score for each sequence *x* was calculated using the temporal change in density at that coordinate as follows:

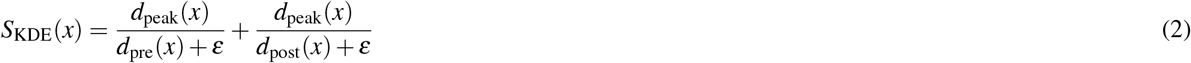

Based on this score, sequences existing in regions where the surrounding density increased rapidly at the peak time point and subsequently decreased were extracted.

## Acknowledgements

The authors thank all study participants and the clinical and technical staff involved in sample collection and repertoire sequencing. This study was supported by JSPS KAKENHI (JP25K08401, JP23K28186), JST FOREST (JPMJFR216J), AMED (JP25ym0126805), and the Uehara Memorial Foundation.

## Author contributions statement

G.M., Y.F., S.I., and M.O. designed the study. G.M., S.I., Y.F., and M.O. developed the methodology. Y.F., K.Y., and G.O. performed investigation. G.M. and S.I. conducted visualization. Y.F., K.Y., H.M., and M.O. acquired funding. Y.F., H.M., and M.O. supervised the project. G.M. wrote the original draft. G.M., S.I., Y.F., K.Y., G.O., H.M., and M.O. reviewed and edited the manuscript. All authors reviewed the manuscript.

## Data availability

The data used in this study consist of human B-cell receptor repertoire sequences derived from hospital patients. These data contain sensitive information and are subject to ethical and legal restrictions approved by the institutional review boards of the participating medical institutions. Therefore, the raw sequencing data and related materials cannot be made publicly available. Access to the data may be considered upon reasonable request, subject to institutional approval and applicable regulations.

